# The SARS-COV-2 outbreak around the Amazon rainforest: the relevance of the airborne transmission

**DOI:** 10.1101/2020.08.06.20169433

**Authors:** Edilson Crema

## Abstract

**Background:** This paper presents a global analysis of the SARS-COV-2 outbreak in Brazil.

Amazonian States have a much higher contamination rate than the southern and southeastern States. So far, no explanation has been provided for this striking difference that can shed light on the airborne transmission of the virus.

Minimizing airborne transmission, health authorities recommend two meters as a safe distance. However, recent experiments reveal that this can be the main form of contagion. There is a lack of theoretical explanation on how airborne transmission works.

**Methods:** To investigate the spread of SARS-COV-2 in different macro environments, we analyzed the daily official data on the evolution of COVID-19 in Brazil. We compared our epidemiologic results obtained in States with very different climatic characteristics, and that had adopted, almost simultaneously, similar social isolation measures. To understand the virus spread, it was necessary to calculate theoretically the movement and behavior in the air of saliva droplets.

**Findings:** The transmission of SARS-COV-2 is much faster in the Amazon rainforest region.

Our theoretical calculations explain and support the empirical results observed in recent experiments that demonstrate the relevance of aerial transmission of the coronavirus.

**Interpretation:** An onset of collective immunity may have been achieved with a contamination rate of about 15% of the Amazonian population. If confirmed, this result will have an essential impact on the management of the pandemic across the planet.

The airborne transmission played a decisive role in the striking difference in the evolution of the pandemic among Brazilian regions.

Air humidity is the most important climatic factor in viral spreading, while usual ambient temperatures do not have strong influence.

There is no safe indoor distance for the coronavirus transmission. So, mask and eye protection are essential.

---

The ray of light that runs through a dark room reveals the existence of numerous small grains of dust that can float in the air for a long time. Since antiquity, this phenomenon was already known. A famous observation of this effect is documented in Lucretius’s poem De Rerum Natura, written around 50 BC. In addition to the empirical description of the phenomenon, and following the tradition of Democritus and Epicurus, Lucretius also proposed an atomistic explanation for the support of particles in the air, according to which their weight would be compensated by the collisions of air atoms.*^1^* However, the behavior of tiny bodies immersed in fluids was only understood from the 19th century on owing to the works of Robert Brown,*^2^* George Gabriel Stokes, and finally with Einstein’s famous work of 1905, *On the movement of small particles in suspension within liquids at rest*.

Currently, this phenomenon has gained tragic relevance due to the uncontrolled dispersion of the Covid-19 throughout the planet, since airborne transmission is one of the forms of viral contamination, as well as the direct reception of drops exhaled by a contaminated person and the contact with infected surfaces. There is still no consensus among researchers as to which of these forms of contagion is the most important in the case of the coronavirus. Despite being the third outbreak of this virus in less than two decades, existing research had not yet fully understood its transmission mechanisms. A similar situation occurred with the Influenza virus. While some important books and works drew attention to the relevance of the transmission by aerosols (droplets)*^3-5^* other authors argued that short-distance transmission by drops would be the main means of infection,*^6,7^* and this latter position prevailed for a long time among health authorities who practically ignored airborne transmission*.^8,9^* At the end of March 2020, the World Health Organization (WHO) released a bulletin stating that there was insufficient scientific evidence that SARS-COV-2 was significantly airborne transmitted. A few months ago, at the beginning of the current pandemic, several governments and the most important health authorities on the planet recommended that only hand washing and a distance of two meters between people would be safe protection procedures and that the use of masks was unnecessary throughout the population. However, with the rapid spread of the coronavirus in countries and in the world, the deadly reality has imposed itself and forced the planetary health authorities to reverse this directive, saving thousands of lives by requiring the use of masks in several countries. From a scientific point of view, this late change in positioning was the authorities’ recognition that air transmission of SARS-COV-2 is an unquestionable fact. Nevertheless, it remains to be understood how this process takes place. In this article we will clarify the physical processes involved in this means of contamination and explain theoretically the results of recent epidemiologic experiments. Besides, we will discuss some recent relevant epidemiologic papers and analyze the SARS-COV-2 outbreak around the Amazon rainforest that may help to understand the relevance of the long-range viral airborne transmission. Finally, we will show that there are still some important recommendations that health authorities should indicate to reduce viral transmissibility.

## Experimental background

The airborne transmission of the coronavirus is now experimentally well demonstrated by important works published during the last months. A relevant study issued in the journal *Nature* revealed the existence of the RNA of the SARS-COV-2 in aerosols collected from the air of several closed environments and open places of two hospitals in Wuhan dedicated only to patients infected with Covid-19 (12).*^10^* Another paper analyzed the air at the Nebraska Hospital Center and also found the SARS-COV-2 in most environments occupied by patients with mild and moderate infections.*^11^* In these two studies it was not possible to confirm if these viruses were active. However, this doubt was finally resolved by a study published in the *New England Journal of Medicine*, where the presence of active SARS-COV-2 in droplets was observed more than three hours after they were artificially produced in the laboratory (65% of relative humidity and temperatures between 21-23°C).*^12^* Now, it is certain that under normal day-to-day conditions SARS-COV-2 remains active for hours in the droplets suspended in the air. Extensive study published in *The Lancet* journal, analyzing empirical data from 16 countries on 6 continents, concluded that the probability of infection by SARS-CoV-2 decreases by 10·6% when using a protection for the eyes.^13^ That is, the risk of contagion through the eyes is very high and continues to be minimized by health authorities, including the WHO, as had happened in the case of masks. This may be a new mistake in combating the pandemic.

On the other hand, further experiments visualized the production of saliva droplets during a person’s normal speech, breathing, sneeze and cough*.^14-16^* Thousands of drops were exhaled and their dispersion in the air was video recorded. They used a laser beam technique of high resolution that was able to identify even submicron droplets. In the video from Kyoto University,*^17^* one can watch the movement of these drops, revealing that while the larger ones fall rapidly and settle on the ground and furniture, there are hundreds of micro droplets that remain suspended in the air several hours after being exhaled. And, most seriously, these small drops disperse rapidly and, a few minutes after their production, occupy the entire environment, covering distances greater than eight meters.

Besides these experiments, there were several empirical situations that put in evidence the dangerousness of the airborne coronavirus infection. Analysis of the most important focus of coronavirus transmission in the 2002-2003 epidemic in Hong Kong, the Amoy Gardens residential complex, demonstrated that a single infected person contaminated, through the ventilation system, more than 300 people living above in the same building. In addition, there are also strong indications that, in the same Amoy Gardens, contaminated droplets could have been carried by the wind for several tens of meters and have infected people in another building, far from patient zero.*^18^* Also, during the coronavirus (MERS-COV) epidemic in South Korea in 2015, research conducted in two hospitals that were sources of contamination revealed that transmission through ambient air may have been one of the main means of contagion.*^19^* Concerning the current outbreak, a recent study demonstrated that the likely onset of the COVID-19 pandemic in Guangzhou, China, occurred by air in a restaurant, where the air conditioning system played a decisive role in the spread of viruses exhaled by an infected man who had just arrived from Wuhan.*^20^* In addition, mass contaminations provoked by only one contaminated person were recently observed in a choral at Los Angeles and a religious cult in France. Finally, a very recent work claims have elucidated the intricate transmission pathways of the new coronavirus and sustains “that the airborne transmission route is highly virulent and dominant for the spread of COVID-19”.*^21^*

## SARS-COV-2 outbreak in the Amazon rainforest

Another new and very relevant epidemiological event has occurred in the current COVID-19 pandemic in Brazil: the Amazonian States that house the forest have presented contamination rates higher than 15%, while in southern States this rate has been less than 1%. To understand this striking difference, we analyzed the official primary data on the pandemic released daily by all States of the country. We will use the number of deaths as an analysis parameter because there is a huge underestimation of the number of cases of the disease due to the extremely low number of tests performed by the Brazilian government. It should be kept in mind that the number of deaths is also undervalued. One of our results is represented in Fig. 1 that shows a tragic consequence of the great difference in the contamination rates, comparing the official number of deaths per week and per million inhabitants between the main Amazonian States and the Southern States of Brazil. The blue arrow in the figure represents the moment of adoption of measures of social isolation by the States.

**Fig. 1.**
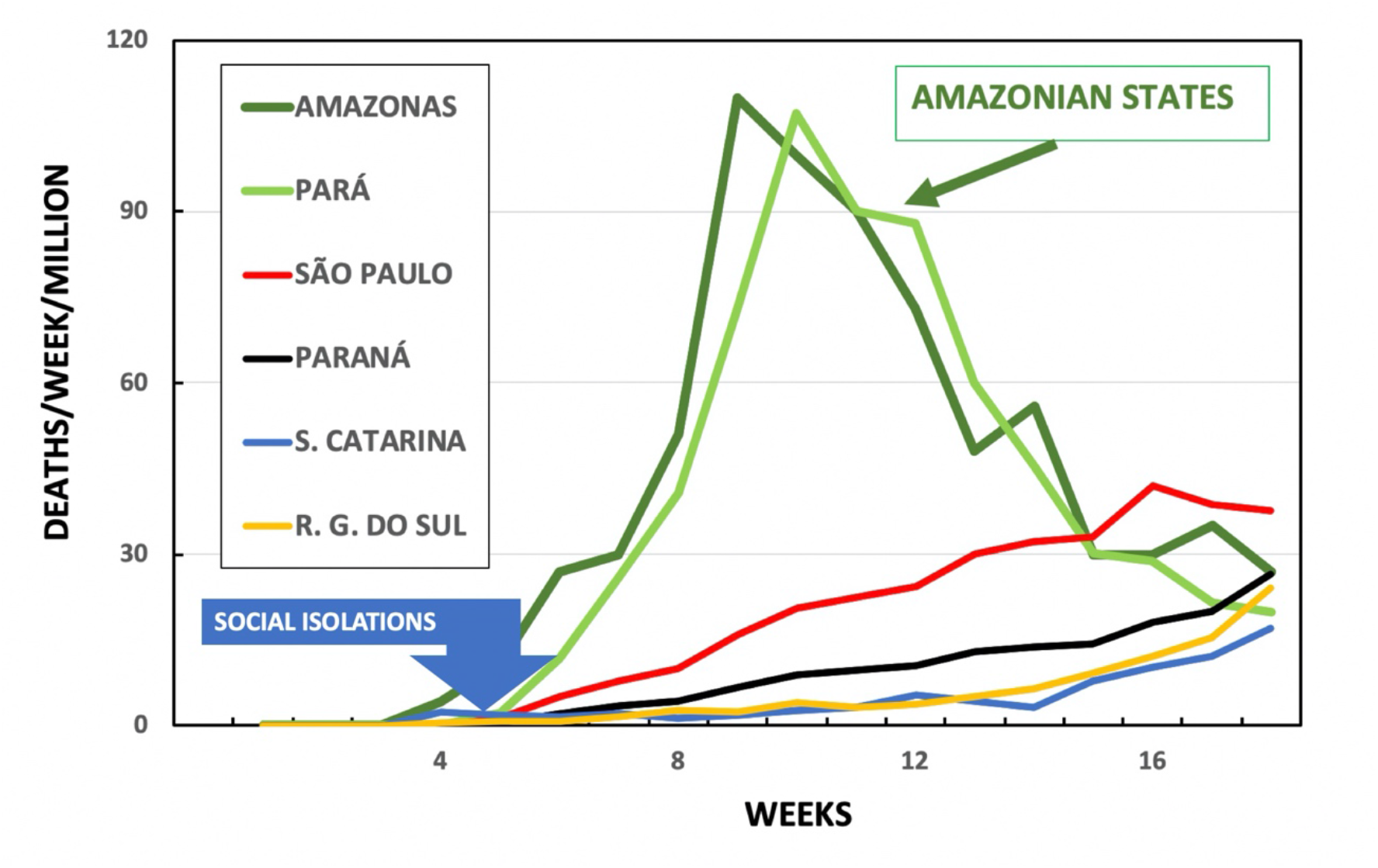
Official number of deaths per week and per million inhabitants during the current SARS-COV-2 epidemic in two Amazonian States and some southern States of Brazil. The large blue arrow indicates the weeks of implementation of social isolation by the States. (Source of primary data: State Health Departments)

These data reveal that near the forest the number of deaths has grown quickly but had also a sharp decrease, while the southern States showed much slower growth. The number of confirmed cases has similar behavior. It is important to emphasize that no additional measures of social isolation were taken after the fifth week indicated in Fig. 1. The evolution of the epidemic in the Amazonian States indicates an extremely important epidemiological result, because the sharp decrease of deaths after reaching the peak was probably due to the large contamination attained in the population. It is surprising that this apparent onset of collective immunity may have been achieved with a much lower percentage of population contamination than predicted by conventional statistical calculations. The actual contamination rate is very difficult to be calculated in Brazil due to the minor number of tests performed. However, extensive serological research conducted by several Brazilian universities calculated that in the State capitals of Amazonas and Pará, during the month of May, the percentage of infected persons was 13% and 15%, respectively. In other cities near the rainforest this percentage attained 20% to 25%.*^22^* Therefore, the contamination rate that could have produced the interruption of the spread of the epidemic observed in Fig. 1 was only of the order of 15%. If confirmed by further studies, this result will have an extraordinary impact on the management of the pandemic on the planet.

The striking difference observed between the north and south regions could not be explained by the issue of social isolation, since both adopted, almost simultaneously, similar isolation measures. Nor is it due to the greater poverty in the northern region, as the southern States also have a large number of inhabitants living in precarious dwellings, including thousands of slums, which would facilitate the rapid spread of the virus. A possible difference in health care system between regions could also not be claimed as an explanation of this phenomenon, as it would not justify the sudden drop in the number of deaths. So, the remarkable disparity in the evolution of the contamination in these regions needs to be deeply investigated.

A factor that can play an important role in the spread of SARS-COV-2 is the climate. During the months investigated in this work, in the regions surrounding the forest, the average temperatures were always above 30°C and the average relative humidity, above 80%. In southern States, average temperatures and average relative humidity were not higher than 22°C and 50%, respectively. Therefore, there is no direct relationship between high ambient temperature and decreased transmissibility. On the other hand, some studies showed that, in general, in environments with relative humidity above 60%, approximately, the drops absorb more than evaporate water into the air.*^23,24^* So, we believe that the airborne transmission of the SARS-COV-2, facilitated by the high humidity of the air, could be a primary factor in the development of the epidemic in Brazil. Our hypothesis is confirmed by other studies conducted in some cities in Brazil at the beginning of the pandemic, although these studies use the number of cases as an analysis variable.*^25,26^* To better understand this relationship, it is interesting to know the amount of water vapor that actually exists in the atmosphere, that is, its absolute humidity. One kilo of air with relative humidity 70%, at 30°C, contains approximately 19g of water in the form of vapor, while at a relative humidity of 40%, at 22°C, the amount of water is only 6g. That is, the process of water evaporation/absorption is very complex in the Amazonian weather conditions, and, on average, the drops expelled by an infected person will absorb water from the atmosphere, allowing the viruses to survive much longer in suspension or deposited on surfaces.

A similar situation has occurred in abattoirs in France, Germany, and the USA that have become huge poles of contamination. The dominant explanation for this phenomenon has been the airborne transmission facilitated by the low temperature of these environments. We agree that the aerial contagion was responsible for the spread of the virus in the abattoirs. However, the pandemic evolution in Brazil, Middle East, Europe, China, and USA has demonstrated that habitual temperatures seem to have little influence on the survival of the virus in the external environment. Therefore, we believe that also in the abattoirs the most important factor was the very high humidity of the air needed in these environments. These examples confirm old and classical work that showed the existence of a very complex relationship between coronavirus survival and air temperature and humidity.*^27^*

Despite all these experimental and empirical epidemiological knowledge accumulated in recent months, there is a lack of a theoretical explanation of the physical behavior of droplets, from their production to their dispersion in the environments. From an epidemiological point of view, the essential physical questions to be answered are how long these infected droplets can remain suspended in the air and what area the droplet cloud can cover in this time. Understanding these physical processes is crucial for health policies-makers, as well as for alerting people about the hidden risks in the environments in which they live.

## Theoretical calculations

During a coughing, sneezing, or speech, thousands of drops of saliva and secretions from the pulmonary tract are expelled with high speeds and penetrate more than two meters into ambient air. These drops have diameters between fractions of micrometers up to fractions of centimeters*.^28-31^* Depending on their composition and air humidity, they can evaporate or absorb water from the environment, and this process is fundamental to the survival of the virus. After the evaporation or growth of the drop, its equilibrium size will define its dynamics and the progression of the epidemic. The smaller droplets (diameter<10 μm) are responsible for the airborne transmission that has been recently recognized by numerous researchers*^32^* as decisive in the spread of the SARS-COV-2 and the WHO asks that it be better studied. In an attempt to contribute to this scientific effort, this work will also investigate the movement of these droplets to demonstrate how airborne transmission is physically possible. Two situations will be considered: they fall into the air at rest; and under action of a vertical airflow upwards which can be produced by an air conditioning system and/or air renewal.

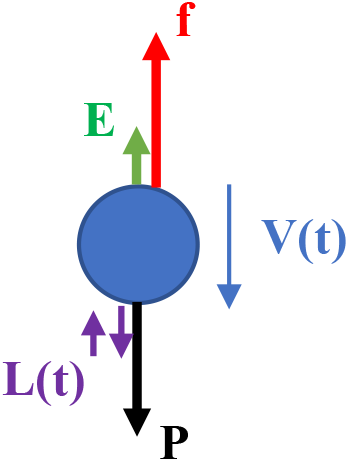

Let us consider the vertical movement of a small spherical drop, with radius R and mass M, immersed in the air, supposed to be homogeneous and in thermal equilibrium at temperature T. The diagram on the side represents the vertical forces acting on the drop: its weight **(P);** the buoyancy **(E)** caused by the air it displaces; the viscous frictional force **(f)** of the air; and, finally, the vertical component of a time-dependent force, **L**(t), extremely complex, caused by the random fluctuation of the collisions of air molecules with the drop, called Langevin force. This last force is significant only for very small particles when statistical variations in air density can cause macroscopic displacements of the drop, the so-called Brownian motion. The differential equation that governs the velocity of the drop’s fall will be:

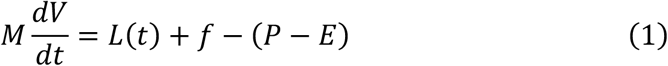

As we are interested in low speeds, the viscous frictional force can be approximated by ƒ = − *μV*, where μ is the viscous friction coefficient of air, given by Stokes’ law, which, in the case of a sphere of radius R, will be *μ =* 6πηR. The air viscosity coefficient, η, depends on the ambient temperature, atmospheric pressure and drop radius. The exact solution of Eq. 1 is very complex due to the random characteristics of the Langevin force. However, when applied to our 0·1 μm radius drops, separately and in one direction, Einstein’s equations give mean square displacements in the position of only 0.1 mm per hour. Larger drops will have even smaller dispersions. That is, we can disregard the Brownian effect in our evaluation. In this case, the solution is very simple, and the drop that starts from rest will fall with the velocity:

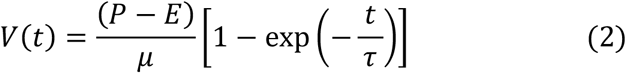

where 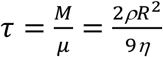 the constant that governs the temporal behavior of the velocity of the drops, and ρ is the density of the substance that constitutes them. Equation 2 shows that their movement is extremely dependent on their sizes. To analyze the epidemiological implications of this expression, we will calculate the fall of drops of human saliva in the air at a pressure of 700 mmHg and a temperature of 25°C.

Table 1 shows the values of the time constant τ calculated for drops of radius ranging from 0·1μm to 50 μm. On the other hand, Fig. 2 shows the dynamics of two drops, of radii 0·5 μm and 1·0 μm, where are shown the variations of their velocities with time. We observed that the drops start to fall from rest, increase their velocities, and in a few microseconds acquire extremely small constant velocities, V_lim_. For example, in this ideal situation, with stationary air, a drop of radius 0·1 μm falls with speed 2·2 μm/s, and a drop of radius 0·5 μm falls with speed 35 μm/s, approximately.

**Table 1.** Fall of saliva droplets with radius R, where τ are their time constants, V_lim_ are their final velocities and T_tot_ are the total time they take to travel the height of 1 · 80 meters.

| $R$ ($\mu\text{m}$) | $\tau$ ($\mu\text{seg}$) | $V_{lim}$ (mm/s) | $T_{tot}$ |
| --- | --- | --- | --- |
| 0.1 | 0.23 | 0.0022 | 227 hours |
| 0.5 | 3.55 | 0.035 | 14.3 hours |
| 1.0 | 13.1 | 0.13 | 3.8 hours |
| 10.0 | $1.3 \times 10^3$ | 11.9 | 2.5 min |
| 50.0 | $3.0 \times 10^4$ | 296.3 | 6.1 seg |

**Figure 2.**
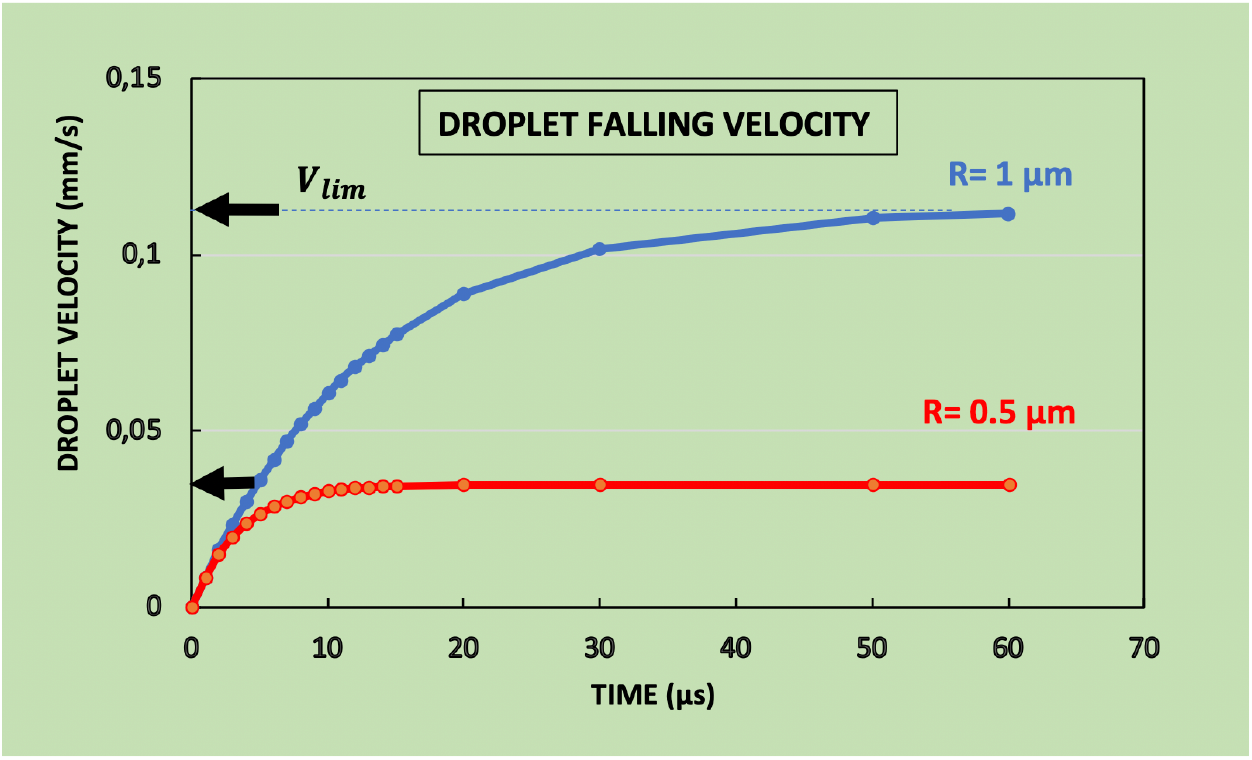
Velocities of two droplets of saliva, radius 0·5 μm and 1·0 μm, which fall into the air from rest.

Evidently, these values are negligible when compared to the speeds of the random air currents that exist in real environments, which are in the order of cm/s, according the measurements by Matthews et al.*^33^* These internal currents are produced by local differences in temperature and pressure, by the movement of objects or people around the environment, and statistical variation of the pressure. It means that the movement of small suspended particles will be governed by these air currents and they can be dragged until 6 meters or more in 5 minutes. So, these currents are responsible for the movement of dust grains observed by Lucretius, and also for the fluctuation and movement of micro drops filmed in the experiments described above. It is important to remember that the coronavirus has a more or less spherical shape with a diameter of the order of nanometers, that is, the drops we dealt with in this work can carry from hundreds to millions of viruses.

## Air conditioners and heating systems

The last column of Table 1 shows the time, T_tot_, that the drops take to reach the ground starting from a height of 1·80 meters. Even considering the air at rest, it shows that drops of diameters 0·2 μm, 1·0 μm, and 2·0 μm would remain suspended for several hours in the air. However, the air currents in indoor environments are even more important when they have an air renewal and/or conditioning system that creates a continuous flow of air in a more or less fixed direction.*^33^* We will evaluate this phenomenon in the case of an aspiration system placed on the ceiling of the environment, producing an airflow in the vertically upward direction, with speed **V**. The diagram on the side shows the forces acting on it. In this case, the viscous friction force of the air passing through the droplet can compensate for and/or overcome the weight force, causing it to remain stopped and/or be aspirated towards the ceiling. Let’s consider the equilibrium limit case, when friction exactly compensates for the weight of the droplet and it stays at rest at a certain height of the ground. Disregarding Langevin’s force, the speed of air that holds a drop at rest can be easily calculated for its different sizes. Figure 3 shows the air velocity necessary to balance drops of different radii, where we observe that they are very small velocities, of a few mm/s, even for the largest drops considered. That is, if the air conditioning system is not well dimensioned, it eliminates the smallest drops, but it can keep larger drops in suspension and/or drastically decrease its fall times, precisely the drops that have a greater potential for infection because they can carry a greater amount of viruses. For example, a continuous vertical flow of air with a speed of only 3 mm/s, approximately, keeps drops of diameter 10 μm suspended at rest for an unlimited time.

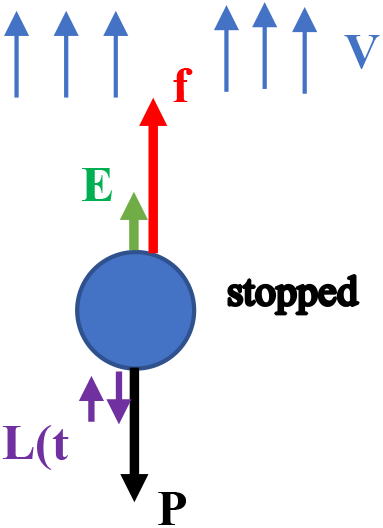

**Figure 3.**
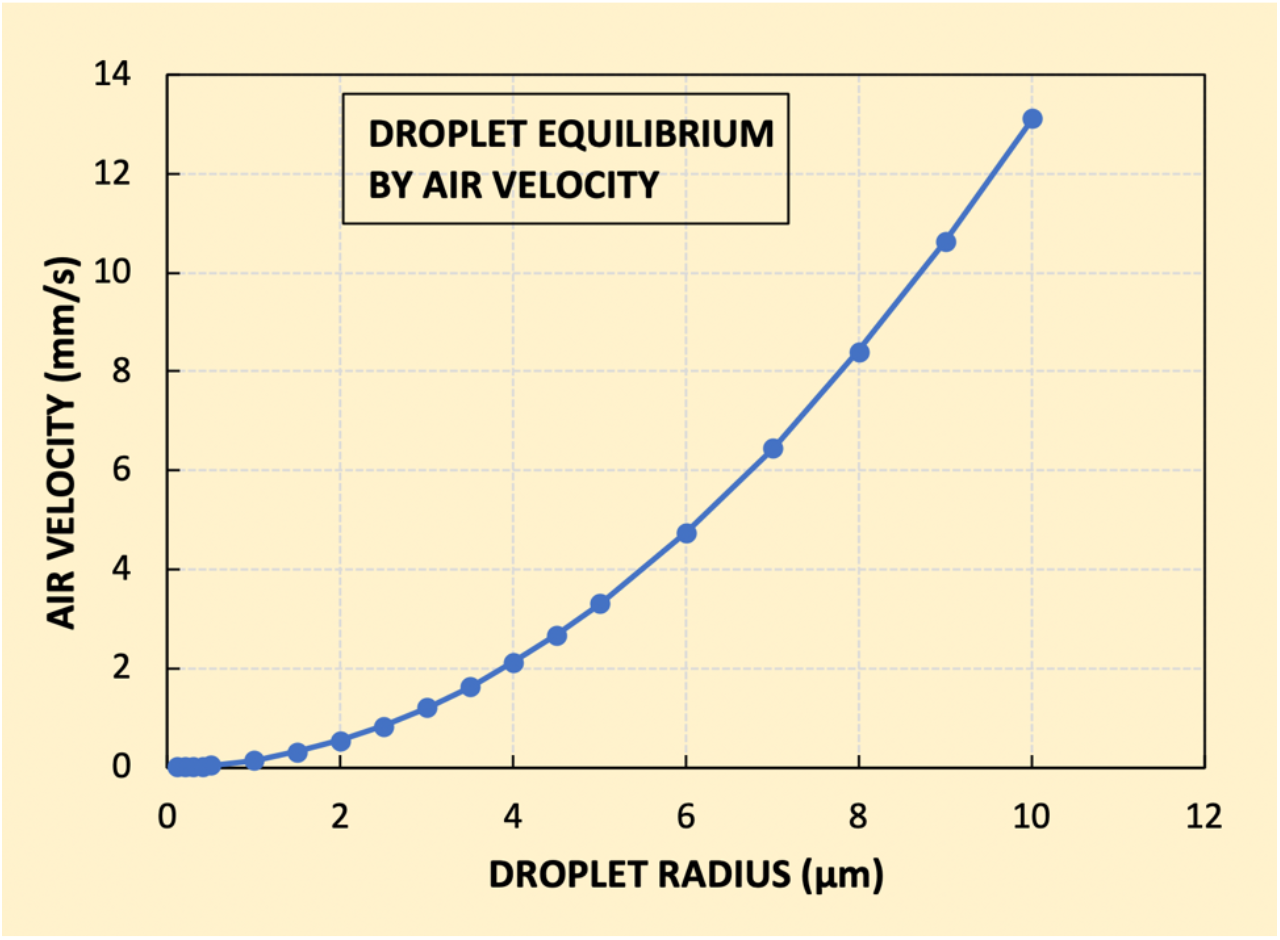
Vertical airflow velocity required to keep drops of saliva suspended in the air at rest.

Finally, we must discuss the effect of heating systems placed on the ground, as they also create an upward airflow and can keep contaminated droplets floating in the air for a long time, increasing the risk of contamination. Perhaps, this may be one of the factors that increase the spread of viral epidemics during the winter.

## Conclusions and warnings

It is now experimentally and theoretically demonstrated that airborne contagion by SARS-COV-2 can occur long after an infected person has spoken, coughed, or sneezed in an environment. These scientific results call into question one of the main recommendations of health authorities to contain the outbreak: the distance of 1m to 2m between people would be a safe method of prevention. This indication is based only on old studies about the direct transmission by larger drops, dangerously ignoring the contamination by the virus airborne in droplets that remain suspended in the air for several hours, and even days after the environment has been visited by an infected person. This recommendation created in the population the false idea that, by staying two meters from each other, it is not necessary to use a mask or other protections. Even the highest leaders of the WHO conduct daily interviews without a mask.

Therefore, an important warning must be made: the distance of two meters is not safe for those who do not wear a mask and, if we consider the infection by the eyes, the distance of two meters is not safe even for those wearing a mask.

The very high SARS-COV-2 transmission rates in Amazonian States in Brazil and many abattoirs around the world provide empirical corroboration of the relevance of the airborne way of contagion. These two environments have high air humidity that allows viruses to survive much longer in droplets in suspension or deposited on surfaces. Therefore, air humidity seems to be the major climatic factor in the development of the COVID-19 epidemic. On the other hand, apparently, there is no direct relationship between high ambient temperature and decreased transmissibility. Besides, the collective immunity in Amazonian States may have been achieved with a contamination rate of around 15% of the population, much lower than predict conventional statistical studies, and which would have an extraordinary impact on pandemic management across the planet.

The argument that the wind disperses the drops has made people feel more protected in open places. However, the same wind that can disperse the drops can also carry them and project them on passersby, whether on the beach, on the street, in the elevator, at home, or on public transport. So, in addition to the mask the use of eye protection should also be recommended. It should not be forgotten that drops in suspension can also be deposited on our face, hair, and clothes.

Finally, another important alert about air conditioning and heating systems comes from our calculations: if they are poorly positioned and/or sized, they can work as a dangerous spreader of viruses.

## Data Availability

No data

